# Adaptive executive control indexed by frontal theta/beta dynamics and trial-level EEG during ecologically valid tasks in schizophrenia

**DOI:** 10.64898/2026.07.08.26357605

**Authors:** Winai Chatthong, Maliwan Rueankam, Supalak Khemthong

**Affiliations:** Assistant Professor of Occupational Therapy, Faculty of Physical Therapy, Division of Occupational Therapy, Mahidol University, Salaya, Nakhon Pathom 73170, Thailand

**Keywords:** schizophrenia, healthy controls, theta/beta ratio, QEEG, executive function, augmented reality, neural biomarkers

## Abstract

**Background:** Schizophrenia is characterized by persistent executive dysfunction and atypical engagement of prefrontal circuits underlying attentional control. However, the neural dynamics of executive processing during ecologically relevant tasks remain underexplored. This study examined frontal theta/beta oscillatory patterns and trial-level EEG responsiveness as indices of adaptive cognitive control in schizophrenia compared to healthy controls.

**Methods:** Thirty adults with schizophrenia (*M* age = 39.9, *SD* = 9.10 years) and a matched healthy control group (*N* = 30; *M* age = 32.25, *SD* = 6.50 years) underwent quantitative EEG during an eyes-open resting state and while performing two executive tasks: an augmented reality visuomotor challenge (LCAR) and a mobile-guided daily routine task (B2B). Frontal theta/beta ratios (TBR) at Fz and Cz indexed attentional engagement. Trial-level responsiveness was assessed via discrete Stimulus–Response Events (SREs).

**Results:** Both LCAR and B2B elicited significant TBR increases relative to eyes-open rest at midline frontal sites (*p* < .001), reflecting elevated executive demand. Compared to healthy controls, participants with schizophrenia exhibited higher baseline TBR and reduced modulation across task segments. In contrast, controls showed stronger SRE-linked variability and greater memory gains, indicating more efficient task-locked cognitive adaptation. Age-related effects were also observed, with participants under 40 years showing higher resting TBR at Fp1 (*p* =.01).

**Conclusion:** Findings advance understanding of prefrontal theta/beta modulation as a neurophysiological marker of adaptive executive control during complex, ecologically valid tasks. By integrating real-world paradigms with trial-level EEG analyses, this study contributes to models of dynamic information processing and cognitive resource allocation in schizophrenia.

## Introduction

Schizophrenia is a chronic, heterogeneous neurodevelopmental disorder characterized by persistent disruptions in executive function (EF), including working memory, cognitive flexibility, inhibition, and goal-directed planning (Cavanagh & Frank, 2014; Kluwe-Schiavon et al., 2013). These deficits strongly predict long-term functional disability, limiting social participation, independent living, and treatment adherence (Synovec, 2015; Tang et al., 2019).

Neurophysiologically, schizophrenia involves dysregulation of frontal cortical networks, marked by aberrant theta oscillations and reduced adaptive engagement of executive circuits highlighting fundamental disruptions in human information processing (Hamedi et al., 2025).

Quantitative electroencephalography (QEEG) provides a noninvasive window into these disruptions. The frontal theta/beta ratio (TBR) is an established neurophysiological marker of top-down attentional control, where increased theta relative to beta power reflects heightened executive demand and reduced regulatory capacity (Cavanagh & Frank, 2014; Knyazev, 2012). This pattern aligns with conflict monitoring models that implicate frontal midline theta in detecting and resolving cognitive interference (Botvinick et al., 2001), and with load theory, which describes how capacity constraints shape adaptive allocation of attentional resources under increasing demands (Lavie et al., 2004). Yet most EEG studies rely on resting-state or block-level analyses, limiting insights into how the brain dynamically reallocates control on a moment-to-moment basis an essential feature of adaptive information processing.

Beyond its theoretical role in models of attentional control, TBR has gained translational significance through evidence of abnormalities across psychiatric disorders. Newson and Thiagarajan (2019) documented disrupted resting-state TBR patterns in schizophrenia and other conditions, underscoring its relevance as a biomarker of cognitive dysregulation. On this basis, the present study employs TBR as a clinically grounded neurophysiological index to examine executive engagement in schizophrenia.

Recent work has advanced ecological and theoretical relevance by embedding cognitive assessments within real-world simulations. For example, the Luk Chup Augmented Reality (LCAR) task integrates sequencing and visuomotor demands within culturally meaningful contexts (Hinz, 2019; Khemthong & Schrepp, 2019), while the Brainy2Blessly (B2B) mobile app captures attention and memory engagement through daily routines (Chatthong et al., 2020a). Coupled with QEEG, these paradigms extend cognitive control models beyond laboratory tasks, illuminating how neural systems adjust to complex, realistic challenges (Bauer et al., 2020; Beauchemin et al., 2024).

Critically, trial-level EEG indices, conceptualized here as Stimulus–Response Events (SREs), enable fine-grained tracking of executive modulation across discrete cognitive events. This approach advances theories of adaptive control by providing empirical resolution into real-time adjustments (Bello et al., 2025; Jia et al., 2021). Moreover, examining age-related differences in these dynamics may illuminate lifespan trajectories of prefrontal efficiency and compensatory mechanisms (Ferguson et al., 2021).

Accordingly, this study aimed to (1) compare within-subject TBR across Eyes-Open baseline and two ecologically embedded tasks (LCAR and B2B), (2) assess how age influences frontal engagement, and (3) identify trial-specific SREs that reflect adaptive shifts in executive load. By situating TBR and trial-level EEG within culturally grounded, naturalistic tasks, this work advances foundational models of dynamic cognitive control and deepens understanding of human information processing in schizophrenia (Csukly et al., 2024).

## Methods

### 2.1 Participants Characteristics and Ethical Approval

Thirty adults diagnosed with schizophrenia (mean age = 39.9 ± 9.10 years; 10 male, 20 female; all right-handed held an undergraduate-level education) participated in this study. Participants were recruited from outpatient psychiatric services and met diagnostic criteria for schizophrenia according to both the DSM-5 and ICD-10 (F20.0). Clinical stability was confirmed using the Thai version of the Positive and Negative Syndrome Scale (PANSS), which has demonstrated acceptable validity and interrater reliability (Nilchaikovit et al., 2000). To ensure scoring consistency, PANSS ratings were independently completed by two trained psychiatric nurses and one psychiatrist serving as the gold standard. Interrater reliability was high across PANSS domains, with ICC values ranging from 0.86 to 0.91. Pearson correlations between nurse ratings and psychiatrist scores demonstrated strong concurrent validity (r = 0.89–0.93), supporting the reliability and accuracy of symptom assessments.

All participants scored in the minimal range across PANSS subscales, consistent with stable clinical status. No participant exceeded a total PANSS score of 30, indicating emotional stability and absence of active psychotic symptoms at the time of assessment. Participants demonstrated minimal symptom severity, with mean PANSS scores indicating clinical remission: Positive symptoms (M = 7.1, SD = 0.3), Negative symptoms (M = 7.0, SD = 0.0), General Psychopathology (M = 16.2, SD = 0.6), and Total PANSS score (M = 30.3, SD = 0.7).

Exclusion criteria included acute psychotic symptoms, comorbid neurological disorders, and current substance use. Participants who were taking medications known to significantly affect arousal or EEG measurements such as clozapine or tricyclic antidepressants were excluded from the study. The majority of participants were maintained on stable regimens of second-generation antipsychotics (e.g., risperidone, olanzapine, aripiprazole), SSRIs, or mood stabilizers that do not confound EEG biomarkers.

Sex was recorded based on binary classifications (male or female) as documented in participants’ medical records; gender identity was not assessed. Given the modest sample size (n = 30), sex- or gender-stratified analyses were not performed.

All participants provided written informed consent. The study protocol received ethical approval from the institutional review boards of [blinded for review].

### 2.2 Experimental Task 1: B2B and Neuroaffective Profiling

The Brainy2Blessly (B2B) protocol is a mobile-based cognitive task designed to assess real-time executive engagement through interactive daily routines. Participants completed two monitored sessions with QEEG recording prior to the B2B task. The monitored session comprised:

- A 3-minute resting condition (eyes open)
- A 3-minute calming period (eyes closed)
- A memory task with four 30-second image recall trials QEEG data were continuously acquired during these conditions.

As part of a neuroaffective profiling module, participants also rated their engagement in 16 real-life activities theoretically linked to four major neuromodulators—dopamine, oxytocin, serotonin, and endorphin—using the DOSE framework (Breuning, 2016). Representative activities included cooking, group singing, journaling, and jogging. Reported frequencies were converted to metabolic equivalent task (MET) values based on the Thai Physical Activity Guideline (2020), allowing for the computation of a normalized engagement score for each neurotransmitter system.

### 2.3 Experimental Task 2: LCAR

The Luk Chup Augmented Reality (LCAR) task is a culturally embedded cognitive challenge that involves AR-guided modeling of traditional Thai fruit-shaped objects, followed by a shape-color memory phase. Delivered through a wearable display, this task engages visuomotor processing, working memory, and attention across six progressively complex object levels (see Table 1).

**Table 1.**
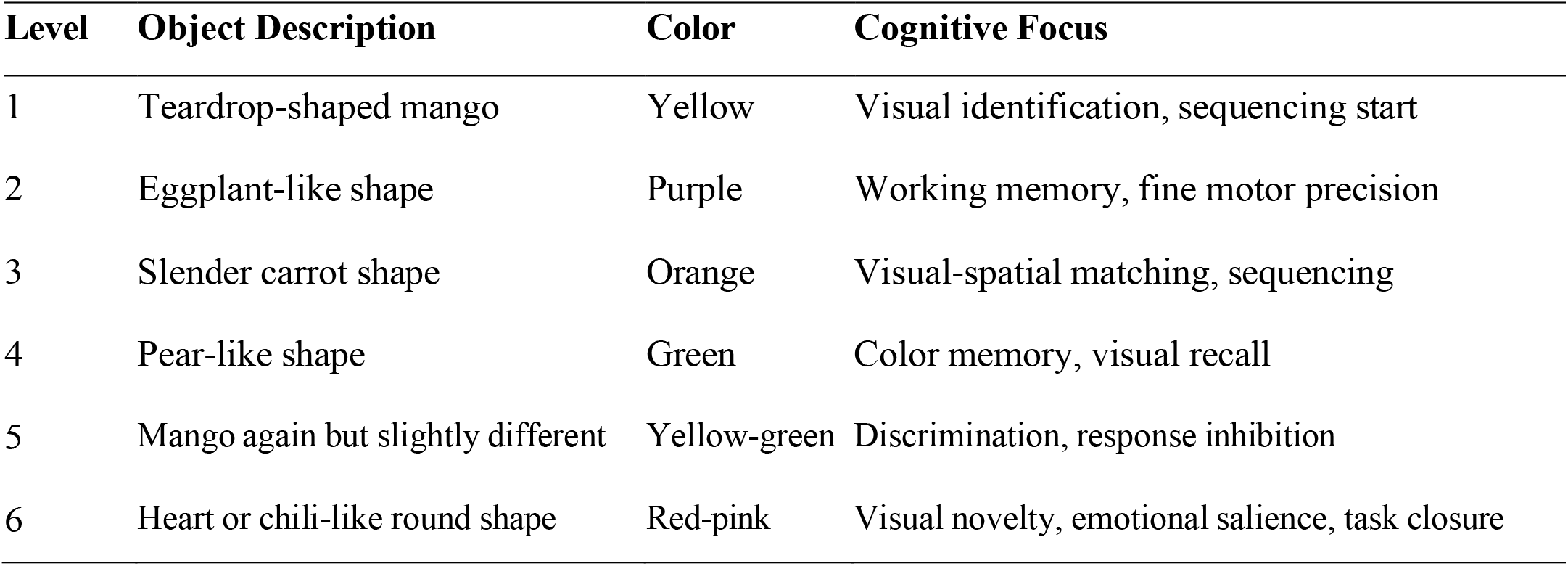
Descriptions of the six sequential LCAR task levels, including modeled clay object characteristics, associated colors, and executive function domains targeted by each level. These levels integrate visual perception, memory recall, and fine motor skills.

| Level | Object Description | Color | Cognitive Focus |
| --- | --- | --- | --- |
| 1 | Teardrop-shaped mango | Yellow | Visual identification, sequencing start |
| 2 | Eggplant-like shape | Purple | Working memory, fine motor precision |
| 3 | Slender carrot shape | Orange | Visual-spatial matching, sequencing |
| 4 | Pear-like shape | Green | Color memory, visual recall |
| 5 | Mango again but slightly different | Yellow-green | Discrimination, response inhibition |
| 6 | Heart or chili-like round shape | Red-pink | Visual novelty, emotional salience, task closure |

Each level imposes distinct shape and color demands designed to elicit variable executive load (e.g., mango: visual identification; carrot: spatial sequencing; pear: color memory). The task draws conceptually from cognitive developmental theory (Inhelder et al., 2014), tactile memory research (Ayres, 1985), and has been validated through user-centered design methods employing the Thai version of the User Experience Questionnaire (Khemthong & Schrepp, 2019). Completion times for each object were recorded as behavioral indices of task efficiency.

### 2.4 QEEG Acquisition and Signal Processing

EEG data were recorded using a 12-channel system arranged according to the international 10–20 system. The active electrodes included Fp1, Fz, F3, F4, F7, F8, Cz, C3, C4, and T3, with linked earlobes (A1–A2) serving as the reference. All electrode impedances were kept below 10 kΩ and continuously monitored. Signals were sampled at 500 Hz, bandpass filtered at 0.5–70 Hz, and a 50 Hz notch filter was applied to minimize line noise. Preprocessing followed guidelines from Keil et al. (2014) and included the following steps:

- Visual inspection for gross artifacts and faulty channels
- Independent Component Analysis (ICA) to remove ocular and muscle-related artifacts (e.g., blinks, eye movements)
- Automated rejection of epochs with voltage deflections exceeding ±100 μV
- Segmentation of data into task-aligned epochs for further analysis

For each condition, power spectral density was calculated using Fast Fourier Transform (FFT) to extract theta (4–7 Hz) and beta (13–30 Hz) band activity. The TBR was computed at each site to reflect dynamic fluctuations in cognitive load and executive engagement. Recordings were obtained across four primary conditions:

1. Eyes-Open baseline
2. Eyes-Closed rest
3. B2B mobile (test and retest for two memory tasks)
4. LCAR (AR-based executive simulation task)

Six regions of interest (Fp1, F7, F4, Fz, Cz, T3) were selected for their relevance to executive function, attentional control, and affective integration. Within-subject variations across experimental conditions and electrode sites were analyzed using repeated-measures ANOVA with Greenhouse–Geisser correction for violations of sphericity.

### 2.5 Trial-Level Engagement via Stimulus-Response Events (SREs)

Stimulus-Response Events (SREs) were defined as temporally discrete EEG segments representing executive activity associated with specific cognitive moments within each task. For each participant, we computed z-scored frontal TBR values aligned to these predefined events, enabling trial-level quantification of attentional engagement and executive modulation (Cavanagh & Frank, 2014; Jia et al., 2021).

To characterize neural engagement with high temporal resolution, we computed 30 Stimulus-Response Events (SREs) based on z-scored TBR values aligned to discrete cognitive events, structured as follows:

- Baseline conditions (SRE_1–2: Eyes Opened, Eyes Closed)
- B2B memory trials (SRE_3–6)
- Neuroaffective prompts (SRE_7–10: DOSE-linked daily activities)
- LCAR modeling levels (SRE_11–16)
- LCAR memory recall levels (SRE_17–22)

This event-related segmentation approach aligns with prior research highlighting the utility of frontal theta/beta dynamics in indexing adaptive engagement across discrete cognitive events (Cavanagh & Frank, 2014; Jia et al., 2021). The SREs were defined as trial-level EEG segments aligned to task-specific cognitive demands. For each trial, the frontal TBR was computed and z-scored relative to each participant’s overall mean.

Single-trial TBR values were then aggregated by condition (e.g., task phase, SRE type) to produce mean SRE scores for statistical analysis. This strategy enabled high temporal resolution during feature extraction, while supporting group-level inference through standardized comparisons.

Within-subject effects were analyzed using repeated-measures ANOVA across conditions and electrode sites. For between-group comparisons, independent-samples t-tests were conducted to evaluate differences in TBR metrics, task performance (e.g., LCAR timing), and trial-level EEG indices (SREs) between participants with schizophrenia (SCZ) and matched healthy controls (HC).

### 2.6 Control Group

A normative healthy control (HC) sample (N = 30; mean age = 32.25 ± 6.50 years) was obtained from an institutional archive of non-clinical EEG participants. HC participants were rigorously screened to ensure the absence of psychiatric or neurological conditions and completed QEEG acquisition, task protocols, and artifact-rejection procedures identical to those employed in the present study. Demographic matching was conducted to closely approximate the age and distributional characteristics of the schizophrenia group, thereby enhancing comparability between groups.

## 3. Results

### 3.1 Frontal TBR Modulation Relative to Baseline

The Eyes-Open resting condition served as the cognitive baseline for evaluating task-related changes in the theta/beta ratio (TBR) at frontal midline sites. TBR values were derived from power spectral densities (PSDs) averaged over theta (4–7 Hz) and beta (13–30 Hz) bands, calculated using fast Fourier transforms (FFT) on artifact-free 2-second epochs. Signal quality was actively monitored, with automatic rejection of segments exceeding ±100 μV or showing muscle artifacts. As summarized in Table 2, ANOVA results indicated a significant effect of task condition on TBR values at Fz, *F*(4, 88) = 13.24, *p* < .001, η^2^ = .38, and at Cz, *F*(4, 88) = 11.07, *p* < .001, η^2^ = .34. Modest increases in theta/beta ratio (TBR) were observed during the eyes closed state relative to eyes-open at both Fz (*M* = 1.59, *SD* = 0.99) and Cz (*M* = 1.38, *SD* = 1.06). However, these differences were not statistically significant (Fz: *p* = .12; Cz: *p* = .28). Topographic maps showed diffuse theta elevation and beta suppression, consistent with inward-focused resting states.

**Table 2.** Theta/Beta Ratio (TBR) at Fz and Cz Across Task Phases Compared to Eyes-Open Baseline.

| Condition | Fz Mean (SD) | F-value | Cz Mean (SD) | F-value |
| --- | --- | --- | --- | --- |
| Eyes Open | 1.36 (0.56) | Baseline | 1.19 (0.60) | Baseline |
| Eyes Closed | 1.59 (0.99) | df (4,88) | 1.38 (1.06) | df (4,88) |
| B2B | 23.27(5.67)** |  | 21.30 (5.04)** |  |
| LCAR - Clay | 1.48 (0.69)* | 13.24, | 1.22 (0.49) | 11.07 |
| Modeling | | $\eta^2 = .38^{**}$ | | $\eta^2 = .34^{**}$ |
| LCAR - Color | 1.28 (0.50) |  | 1.17 (0.47) |  |
| Memory |  |  |  |  |

Note. TBR = Theta/Beta Ratio; SD = Standard Deviation; $p < .05$ (\*), $< .001$ (\*\*); ns = not significant.

In contrast, TBR increased significantly during the B2B task (test phase) at both sites (Fz: *M* = 23.27, *SD* = 5.67; Cz: *M* = 21.30, *SD* = 5.04; both *p* < .001). These elevations were primarily driven by sustained theta activity in the absence of beta enhancement, indicative of heightened executive load and cognitive control. Scalp distributions confirmed midline-frontal theta dominance during B2B, consistent with robust recruitment of frontocortical regulation networks.

During the LCAR Clay Modeling segment, TBR at Fz rose significantly compared to baseline (Fz: *M* = 2.01, *SD* = 1.19; *p* = .05), reflecting increased frontal engagement during integrated visuomotor and working memory tasks. No significant change was found at Cz (*M* = 1.29, *SD* = 1.01; *p* = .71). The subsequent LCAR Color-Memory task did not elicit further significant TBR modulation.

Pairwise comparisons were conducted using Tukey’s HSD. The results confirmed that the B2B task evoked significantly higher TBR values compared to all other conditions at both Fz and Cz sites (*p* < .001). No other pairwise comparisons reached significance, except for the Fz increase during LCAR Clay Modeling (*p* = .05).

### 3.2 Task-Evoked Frontal Engagement and Trial-Level Patterns

A repeated-measures ANOVA revealed significant TBR increases from Eyes-Open baseline to both LCAR and B2B conditions at Fz and Cz, with the strongest effect during B2B. These within-subject differences underscore the elevated executive engagement induced by interactive, augmented, and mobile task environments. Greenhouse–Geisser corrections were applied to adjust for sphericity violations.

Trial-level analyses of z-scored TBR across the 22 Stimulus-Response Events (SREs) indicated stable engagement throughout the protocol. Based on the current dataset (22 SRE conditions across 30 participants), the highest average trial response occurred at SRE_12, with a mean z-scored TBR value of *M* = −0.29. No individual SRE exceeded ±3 SD, suggesting consistent modulation of executive resources across discrete cognitive episodes. This temporal fidelity supports the ecological integration and sequential continuity of the combined task design.

### 3.3 Age-Related Differences in Frontal TBR and SRE Engagement

Comparisons by age group revealed that participants under 40 years exhibited significantly higher resting-state TBR at Fp1 during Eyes-Open baseline (p = .009), indicative of greater preparatory attentional readiness. No significant group differences were found at other frontal (Fz, F4, F7) or temporal (T3, Cz) sites. Exploratory analyses of SREs did not yield statistically significant age effects overall. However, SRE_10—associated with oxytocin-linked B2B activities—approached significance (*p* = .06), with younger participants showing elevated engagement (*M* = 0.37) compared to older participants (*M* = –0.25). Similar nonsignificant trends were noted for SRE_9 (dopamine-related) and SRE_11 (LCAR onset). These patterns suggest potential developmental modulation of task-specific executive responsiveness, warranting further investigation in larger, more age-diverse samples.

### 3.4 Between-Group Differences: Schizophrenia vs. Healthy Controls

Independent-samples t-tests were conducted to compare TBR indices and trial-level EEG markers between the schizophrenia (SCZ) group and a matched normative healthy control (HC) group. Significant group differences were found in frontal theta/beta activity across rest, task, and recall conditions, with medium-to-large effect sizes. Similarly, trial-level analysis revealed attenuated modulation of SREs in the SCZ group across both baseline and task-embedded events. Table 3 summarizes a selection of significant group-level differences in TBR and SRE responses.

**Table 3.** Group Comparisons on Key QEEG and SRE Measures.

| Measure | SCZ Mean (SD) | HC Mean (SD) | t-value | p-value | Cohen's d |
| --- | --- | --- | --- | --- | --- |
| TBECFp1 | 1.61 (1.01) | 1.01 (0.87) | 2.46 | 0.0169 | 0.64 |
| TBECF7 | 1.35 (0.81) | 0.91 (0.68) | 2.32 | 0.0240 | 0.60 |
| TBECFz | 1.59 (0.99) | 1.05 (0.77) | 2.33 | 0.0234 | 0.60 |
| TBEOFp1 | 1.22 (0.48) | 0.81 (0.32) | 3.81 | 0.0004 | 0.98 |
| TBEOFz | 1.36 (0.56) | 1.10 (0.44) | 1.96 | 0.0545 | 0.51 |
| TBBFz | 23.27 (5.67) | 19.52 (4.59) | 2.82 | 0.0067 | 0.73 |
| SRE_1 | -0.06 (0.93) | -0.58 (0.75) | 2.40 | 0.0200 | 0.62 |
| SRE_6 | -0.10 (0.93) | -0.69 (0.71) | 2.75 | 0.0081 | 0.71 |
| SRE_10 | 0.06 (0.93) | -0.49 (0.80) | 2.44 | 0.0181 | 0.63 |
| SRE_17 | 0.09 (1.03) | -0.54 (0.78) | 2.66 | 0.0104 | 0.69 |

To enhance clarity, task-related changes in frontal TBR are also illustrated in Figure 1. The bar graph summarizes mean values at Fz and Cz across all task phases relative to eyes-open baseline and complements Table 3 by highlighting the pronounced elevation during B2B and the more modest modulation during LCAR segments. Figure 1 summarizes group-level TBR differences across six frontal-central EEG electrode sites. The SCZ group consistently showed elevated theta/beta ratios compared to healthy controls, particularly at Fp1, Fz, and T3, indicating greater executive demand and frontal dysregulation.

**Figure 1.**
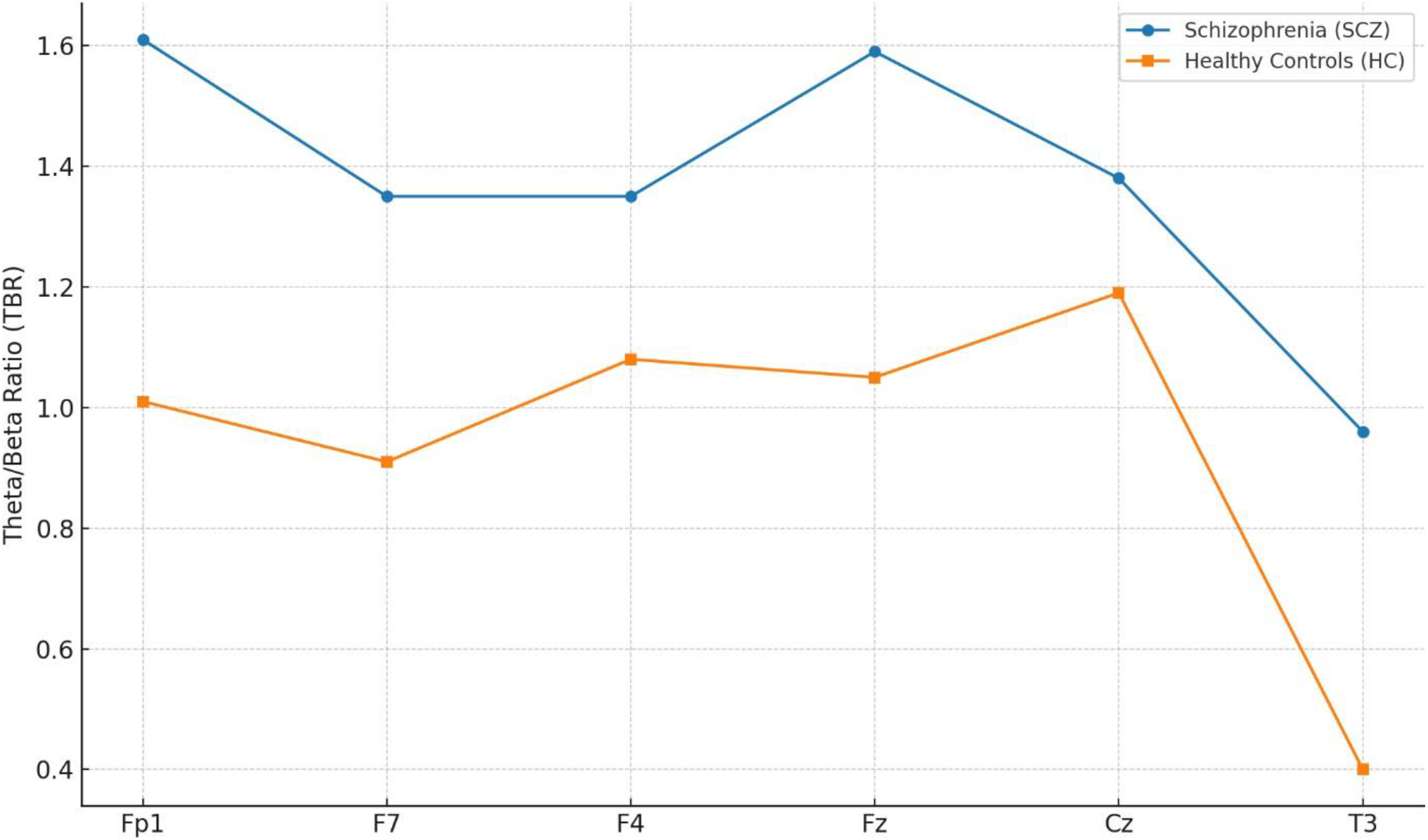
Theta/beta ratio (TBR) comparisons between individuals with schizophrenia (SCZ) and healthy controls (HC) across six frontal–central EEG electrode sites (Fp1, Fz, Cz, F7, F4, and T3). Bars represent group means with standard errors. SCZ participants exhibited consistently elevated TBR, particularly at Fp1, Fz, and T3, indicating greater executive load and dysregulation relative to HC. *HC = healthy controls; SCZ = schizophrenia participants; TBR = theta/beta ratio*.

## 4. Discussion

This study investigated task-evoked modulation of the frontal theta/beta ratio (TBR) and trial-level EEG responsiveness during ecologically relevant executive tasks in individuals with schizophrenia. By integrating quantitative EEG (QEEG) with culturally embedded cognitive challenges, our findings advance theoretical models of adaptive executive control and highlight the temporal dynamics of information processing under varying cognitive demands.

### 4.1 Frontal Theta/Beta an Index of Executive Load and Attentional Allocation

Elevations in frontal theta relative to beta power at Fz and Cz, particularly under cognitive load, align with frameworks positing frontal midline theta (FMT) as a neural marker of effortful control and conflict monitoring (Cavanagh & Frank, 2014; Botvinick et al., 2001). The reduction of theta during eyes-open baseline and its enhancement during task performance support load-sensitive recruitment of medial prefrontal circuits consistent with capacity models of attention and working memory (Tang et al., 2019). Marked TBR increases during both LCAR and B2B tasks further underscore its sensitivity to top-down attentional engagement. Robust elevations during B2B, a mobile task requiring sustained focus, corroborate theories linking frontal theta amplification to goal maintenance and beta desynchronization to reduced motor interference (Knyazev, 2012). Collectively, these patterns reinforce TBR as a fundamental neural index of adaptive adjustments in executive demand and attentional resource allocation.

### 4.2 Regional Differentiation in Cognitive Control

Topographic analyses indicated functional specificity across frontal sites. Fz and Cz were most responsive to global executive load, consistent with anterior cingulate and dorsomedial prefrontal involvement in monitoring and control adjustment (Cavanagh & Frank, 2014; Botvinick et al., 2001; Hamedi et al., 2025). In contrast, lateral regions (F7, T3) may support multimodal or affective integration, while Fp1 likely indexes alertness and preparatory attention. This spatial dissociation advances models distinguishing core control hubs from auxiliary integrative networks (Ferguson et al., 2021; Jia et al., 2021).

A central contribution of this study lies in the trial-level analysis of executive control using z-scored theta/beta ratio (TBR) aligned to Stimulus–Response Events (SREs). This fine-grained approach captured moment-to-moment fluctuations in frontal EEG activity during task execution, revealing how the brain dynamically regulates cognitive effort. Significant correlations emerged between SRE-specific TBR and memory gains, particularly within the Brainy (SRE 3–6) and Blessly (SRE 7–10) segments (see Supplementary Table 4). Elevated TBR during these trials was positively associated with visuospatial memory improvement (r = .81, p < .001) and event-based memory recall (r = .41, p = .10), suggesting adaptive recruitment of attentional and encoding mechanisms.

Group comparisons further revealed that individuals with schizophrenia (SCZ) exhibited elevated baseline TBR and reduced modulation across task segments, consistent with executive control impairments. In contrast, healthy controls (HC) showed stronger SRE-linked variability and greater memory gains, reflecting more efficient task-locked adaptation. Notably, SRE 20 (within the LCAR Color segment) showed peak correlations with memory gain, highlighting specific moments of neurocognitive adjustment where engagement was most efficient (Altman et al., 2023).

Overall, this SRE-based segmentation approach provides empirical support for the utility of momentary EEG metrics particularly TBR as indicators of transient cognitive readiness and learning-related neuroplasticity (Qi et al., 2025; Tang et al., 2019). Figure 2 illustrates mean z-scored TBR across grouped SREs for SCZ and HC participants, highlighting group-level differences in neurophysiological engagement across baseline, memory, and affective segments.

**Figure 2.**
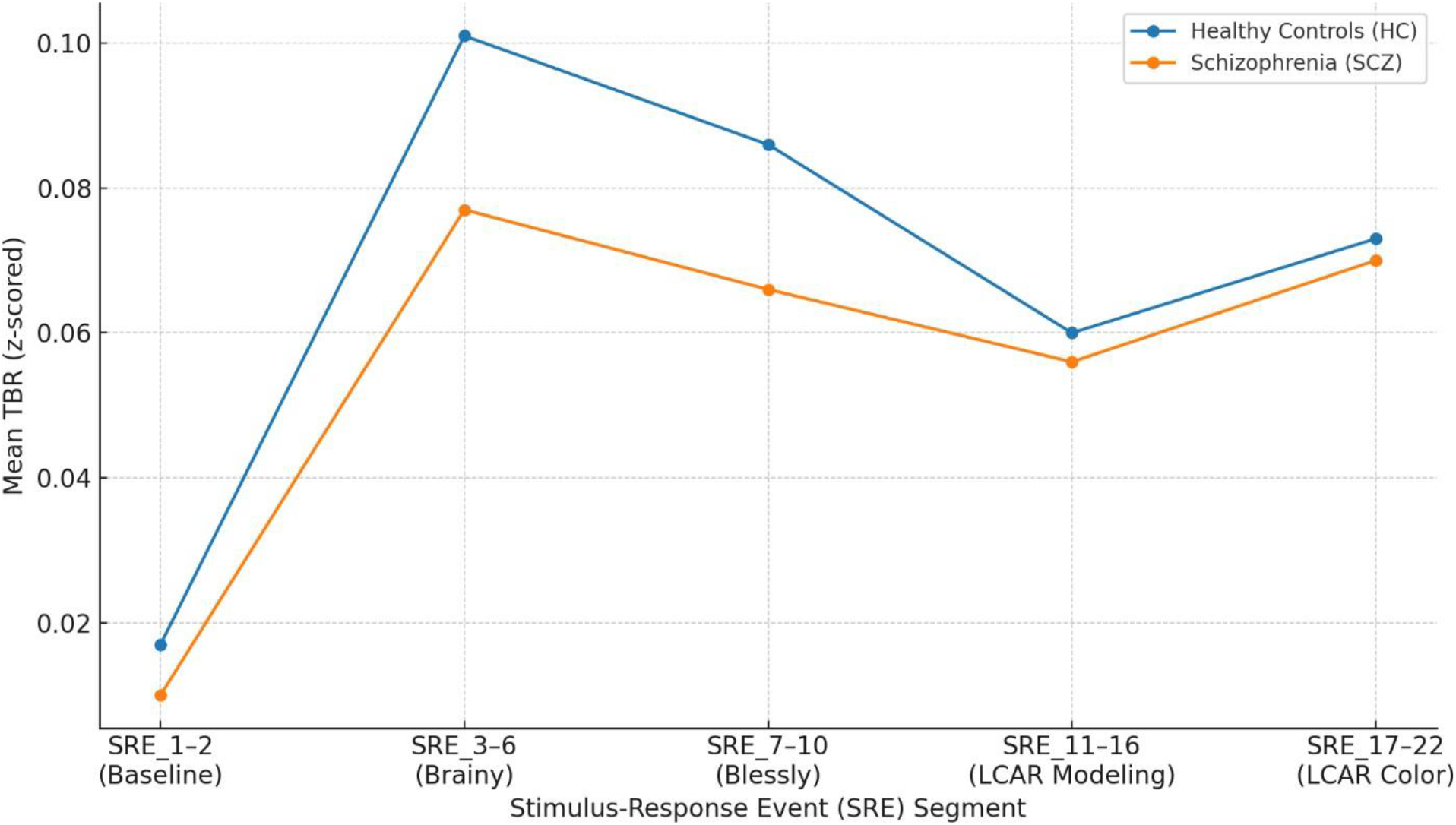
Mean z-scored theta/beta ratio (TBR) across grouped Stimulus–Response Events (SREs) for schizophrenia (SCZ) and healthy control (HC) participants. Bars represent group means with standard errors. SRE segments were categorized as follows: Baseline (SRE 1–2), Brainy task (SRE 3–6), Blessly task (SRE 7–10), Locally Cued Associative Recall [LCAR] Modeling (SRE 11–16), and LCAR Color (SRE 17–22). Compared to HC, SCZ participants showed reduced adaptive modulation of frontal TBR across task segments. *HC = healthy controls; SCZ = schizophrenia; TBR = theta/beta ratio; SRE = Stimulus–Response Event*.

### 4.3 Developmental Modulation of Control

Age-related findings—where participants under 40 showed higher baseline TBR at Fp1 and stronger engagement during social-affective tasks—align with developmental models of prefrontal maturation and compensatory strategies. These results suggest that younger adults may rely on heightened tonic alertness to meet complex demands, underscoring the importance of considering lifespan trajectories in executive control (Ferguson et al., 2021).

### 4.4 Ecological Implications for Executive Function

Embedding executive challenges in culturally meaningful, real-world tasks bridges traditional laboratory paradigms with ecological validity. This design extends information processing models beyond simplified stimuli to contexts resembling everyday cognitive demands. Coupling augmented reality and mobile assessments with trial-level EEG supports emerging perspectives on adaptive, context-sensitive control (Chatthong et al., 2020a; Bauer et al., 2020; Chicchi Giglioli et al., 2019).

### 4.5 Limitations and Future Directions

The modest sample size (n = 30) constrains statistical power and limits generalizability, particularly for detecting subtle group differences or interaction effects. Nonetheless, significant within-subject effects and topographical specificity in TBR dynamics suggest the robustness of trial-level EEG markers. Given the heterogeneity of schizophrenia, these findings should be regarded as preliminary and hypothesis-generating.

Future research should replicate and extend these results in larger, more demographically diverse cohorts that vary in age, illness duration, and medication profiles. Multicenter studies could establish normative benchmarks for SRE-based EEG indices, strengthening their external validity as markers of executive function across real-world contexts.

Another limitation is the absence of gender identity data. Without this, potential neurocognitive differences among gender-diverse individuals cannot be addressed. Future studies should adopt inclusive, self-reported measures of sex and gender in alignment with best practices in sex- and gender-based analysis (SGBA), thereby improving generalizability and supporting equity in cognitive neuroscience.

## Conclusion

This study advances models of executive control by showing that frontal theta/beta dynamics and trial-level EEG markers sensitively index adaptive engagement during ecologically valid tasks in schizophrenia. Significant within-subject TBR differences, age-related variations in attentional readiness, and micro-temporal EEG correlates collectively deepen our understanding of how the brain allocates control under complex, culturally embedded demands. Moving beyond static assessments to capture moment-to-moment neural adjustments, this work contributes foundational insights into human information processing, spanning conflict monitoring, capacity allocation, and developmental modulation.

## Data Availability

All data produced in the present study are available upon reasonable request to the authors

## Supplementary Materials

Supplementary Table 4. Group-Level Theta/Beta Ratio (TBR) Across Task-Aligned Stimulus-Response Events (SRE_1–SRE_22), Memory Gain Indices, and Correlations by Diagnostic Group

